# Basal forebrain volume and concurrent hallucinations and mild cognitive impairment in Parkinson’s disease

**DOI:** 10.1101/2024.12.13.24318999

**Authors:** Sabrina M. Adamo, Vaishali Mutreja, Marta Statucka, Batu Kaya, Taylor W. Schmitz, Anthony C. Ruocco, Melanie Cohn

## Abstract

**Background:** Visual hallucinations (VH) and mild cognitive impairment (MCI) often co-occur in Parkinson’s Disease (PD). Each symptom in isolation is associated with cholinergic basal forebrain (BF) atrophy. However, it is unclear whether BF integrity relates to one symptom preferentially or to their co-occurrence, and whether this relationship is specific to the nucleus basalis of Meynert (Ch4 & Ch4p) or extends to other BF nuclei (Ch1-2 & Ch3).

**Objectives:** To bridge the literature on MCI, VH, and the BF by examining associations between total and subregional BF volumes with VH, MCI, and concurrent symptomology.

**Methods:** We used clinical and structural MRI data in PD patients (n=296), evaluated prior to deep brain stimulation. PD-MCI was identified in 61% of patients, VH in 25%, and symptoms co-occurred in 21% of the sample. We conducted logistic regressions to examine the relationships between BF volumes (total BF, Ch1-2, Ch3, Ch4, and Ch4p) with MCI, VH, and their combination, accounting for potential confounders (age, sex, LEDD, dopamine agonist, and anticholinergic medications).

**Results:** Total BF, Ch1-2 and Ch4 volumes are negatively associated with the likelihood of PD-MCI, but not VH when the other symptom type is not considered. When both symptoms are considered, Total BF and Ch4 volumes are negatively related to the co-occurrence of MCI and VH relative to neither symptom.

**Conclusions:** Our data support the “cholinergic phenotype” in that BF atrophy, including Ch4, is associated with concomitant MCI and VH. Ch1-2 volume is further associated with MCI, which is interesting given its temporal lobe projections.

---

Non-motor symptoms such as visual hallucinations (VH) and mild cognitive impairment (MCI) are common in Parkinson’s disease (PD) [1,2]. Each symptom is a risk factor for the other and for the development of dementia, and their combination may reflect a malignant course associated with a rapid progression towards dementia [3]. While the relationship between VH and MCI is not yet fully understood, cholinergic depletion stemming from basal forebrain (BF) degeneration may be a key contributor to each symptom. Pasquini et al. [4] proposed a “cholinergic phenotype” comprising the presence of MCI, VH, as well as other symptoms (e.g., gait problems, sleep disturbances, and olfactory impairment). Importantly, this proposal is based on their review of neuroimaging (PET, SPECT, & MRI) and post-mortem pathology studies showing cholinergic and BF alterations in PD-VH and PD-MCI in isolation, rather than in patients with concomitant symptomatology, as studies rarely report the co-occurrence of these symptoms. Thus, it remains uncertain whether BF alterations and associated cholinergic loss are preferentially associated with one symptom type or with concomitant symptoms, as their frequent co-occurrence may have created a confounding or third variable problem in previous research.

The BF, which is the main sources of acetylcholine to the brain, is comprised of multiple cholinergic nuclei. Ch1-2 project mainly to the hippocampi, Ch3 projects to the olfactory bulb and entorhinal cortex, and the nucleus basalis of Meynert (NBM), which can be segmented into anterior (Ch4) and posterior (Ch4p) regions, projects to the neocortex and basolateral amygdala [5,6]. NBM alterations are associated with VH [7] and with cognitive decline [8,9] in PD. In addition, reduced white matter integrity of BF projections has been shown in PD-VH compared with PD-noVH and controls groups [10], and in the NBM of PD-MCI patients compared with PD patients with intact cognition [8]. Post-mortem pathology studies showed a buildup of Lewy bodies in the NBM of PD patients with well-formed VH as well as with PD dementia [11]. To date, there is limited research on the other anterior BF nuclei (Ch1-2-3), and of those limited studies, the reported results either pertain to Ch1-2 alone or in combination with Ch3, while Ch3 has not been assessed on its own [6]. Specifically, only three studies examine these anterior BF nuclei in PD [7,9,12]; none of which report volumetric loss in PD-MCI or PD-VH versus their cognitively intact or non-hallucinating counterparts [7,9], and only one study found differences in Ch1-2-3 and Ch4 between PD patients and healthy controls [12], but with no report of whether patients presented with VH or MCI.

Importantly, no studies have tested the cholinergic phenotype proposal directly by investigating BF structural integrity in patients with concomitant PD-MCI and VH. Furthermore, research on individual symptom type focuses primarily on the Ch4 within a single early PD dataset (i.e., Parkinson’s Progression Markers Initiative; [7–9], but see [12,13] for other cohorts). This literature is further muddied by the confounding effects of medications [levodopa, dopamine agonists (DA), and anticholinergic medications], which may, at high dose, promote or exacerbate both cognitive [14,15] and VH symptoms [15,16].

The aim of this study is to test the cholinergic phenotype proposed by Pasquini et al. [4], and by doing so, bridge the two bodies of literature on VH, MCI, and the BF. Specifically, our goal is to examine whether the BF is a common neural substrate of PD-VH and PD-MCI while accounting for the confounding effects of demographic characteristics and both dopaminergic and anticholinergic pharmacotherapy. To achieve this, we performed a retrospective clinical chart review consisting of data from a multidisciplinary assessment (neuropsychology, neurology) and structural MRI completed prior to Deep Brain Stimulation (DBS). First, we hypothesize that total BF volume, and specifically the NBM, is associated with MCI as well as VH when examined in isolation, as suggested by the literature on each symptom type. Second, based on the cholinergic phenotype proposal suggesting that VH and MCI have a compounding effect, we hypothesize that smaller total BF and NBM volumes are associated with concomitant symptoms relative to groups presenting with one symptom or neither. As the other BF nuclei (Ch1-2, Ch3) have seldom been investigated [6], we explore whether similar patterns are found with these regions.

## METHODS

### Participants

We conducted a chart review of PD patients who underwent multidisciplinary assessments and 3T MRI prior to DBS at the Toronto Western Hospital between January 2007 and August 2021. Exclusion criteria were: poor imaging quality, > six-month gap between the neuropsychological assessment and MRI, age < 40, neurological disorders other than PD (e.g. stroke, epilepsy, moderate to severe traumatic brain injury), severe mental illness (schizophrenia, personality disorders), severe scores on depression scales [Beck Depression Inventory-II (BDI-II) > 28, or Geriatric Depression Scale (GDS) > 19)]; [17,18], PD dementia based on Emre et al. 2007 criteria [19] and dementia with Lewy bodies based on McKeith et al. 2017 criteria [20], limited English fluency, level I neuropsychological assessment [21], and missing VH data. A total of 296 people met inclusion criteria.

### Sample characteristics

Demographic variables included age, sex, and years of education. Clinical variables pertaining to pharmacotherapy included levodopa equivalent daily dose (LEDD) [22], whether patients took DA (yes/no), and whether they are prescribed high levels of anticholinergic medications (i.e., score ≥ 3 on the Anticholinergic Cognitive Burden Scale (ACB) [23]). Specific DA and anticholinergic medications, including antipsychotics and cognitive enhancers, are reported in Table S1. Additional clinical variables included disease duration (based on diagnosis date), motor symptom severity ON and OFF [Movement Disorders Society-Unified Parkinson’s Disease Rating Scale Part III (MDS-UPDRS-III)] [24], and percent levodopa response [(OFF– ON)/OFF*100%]. UPDRS-III [25] scores were converted to MDS-UPDRS-III scores for 218 patients using a validated transformation method [26]. OFF scores were not available for seven patients who did not complete a levodopa challenge within six months of the MRI or neuropsychological testing.

### Visual Hallucination Status

All patients completed a semi-structured clinical interview alone (14%) or with their care-partners (86%) during which they were asked whether they experience minor VH (feelings of presence, passage illusions, visual misperceptions), well-formed hallucinations or delusions, and whether they retained insight vis-à-vis these experiences. These open-ended descriptions were summarized in the clinical neuropsychological reports and the presence and severity of VH were coded on the MDS-UPDRS [24] item 1.2 by two independent raters with high interrater reliability (Cohen’s Kappa>0.8), and discrepancies were resolved by consensus [22] This item ranges from 0-4 where 0=absence of hallucinations, 1=minor hallucinations, 2=well-formed hallucinations with preserved insight, 3=well-formed hallucinations with loss of insight, and 4=delusions. Scores were then binarized (yes/no), and patients with a score > 0 were included in the VH group. More fine-grained severity ratings and subtypes of minor hallucinations are reported in Table S2.

### PD-MCI status

Cognitive diagnoses (PD-MCI versus intact cognition) were based on a level II neuropsychological assessment [21] and reached via consensus by licenced neuropsychologists (M.S. and M.C.) based on the presence of subjective cognitive decline, preserved independence with instrumental activities of daily living, and performance falling 1.5 SD below normative data on at least two independent neuropsychological tests. Visuoperceptual and spatial measures included the Benton Judgement of Line Orientation [27], Object Decision and Silhouette subtests from the Visual Object and Space Perception battery [28], and the Rey-Osterrieth (Rey-O) Figure Copy test [29]. Language tests comprised of the Boston Naming Test [30], Vocabulary subtest from the Wechsler Abbreviated Scale of Intelligence (WASI or WASI-II) [31,32], and the Category Verbal Fluency subtest from the Delis-Kaplan Executive Function System (D-KEFS) [33]. Memory tests included the California Verbal Learning Test-II [34], Logical Memory I and II subtests from the Wechsler Memory Scale-III (WMS-III) [35], and the Rey-O recognition test [29]. Attention and processing speed were assessed using the Trail Making Test-A (TMT-A) [36], D-KEFS Stroop colour and reading speed [33], and the Digit Span subtest from the WMS-III [35]. Lastly, executive functioning was assessed using the D-KEFS Letter and Switching Verbal Fluency, D-KEFS Stroop interference and switching [33], TMT-B [36], Wisconsin Card Sorting Test [37], Matrix Reasoning subtest from the WASI or WASI-II [31,32], and the Conditional Associative Learning Test [38]. The BDI-II [17] or GDS [18] questionnaires were also collected and used for the exclusion criteria. MCI subtypes and cognitive domains affected are described in Table S3.

### MRI acquisition and preprocessing

T1-weighted spoiled gradient recalled echo (SPGR) structural MRI scans were acquired on a 3T Signa MR system (GE Medical Systems, Milwaukee, WI). Images were resliced to 1×1×1mm voxel dimensions due to some variability in acquisition parameters (voxels acquired at <1mm^3^, see Table S4). MRIQC [39] was used for visual inspection. A voxel-based morphometry pipeline previously used in Alzheimer’s disease [40,41] was implemented in SPM12 (http://www.fil.ion.ucl.ac.uk/spm) and MATLAB 2018a. First, images were bias-corrected to adjust for inhomogeneity and aligned into anterior/posterior commissure space before being affine co-registered to the Montreal Neurological Institute (MNI) template space. Grey matter (GM), white matter, cerebrospinal fluid, and total intracranial volume (TIV) segmentations were then generated on the T1 images and moved to a study-specific template using DARTEL. A study-specific template was used to minimize the geodesic distance from each subject to the population mean [42]. Non-linear modulation of GM images was done using the CAT12 toolbox, resulting in voxel-based indices of GM volume. There was no spatial smoothing applied to the volumes. Scans with poor imaging quality based on visual inspection or ones that failed CAT12 quality pipeline (e.g., homogeneity outliers) were excluded (n=4). Four bilateral BF ROIs (Ch1-2, Ch3, Ch4, Ch4p) reflecting different BF subregions derived from histological post-mortem studies [43] were segmented using a threshold of 50% (which is optimal for BF volume extraction [44]). As these ROIs are in MNI space, they were warped to the DARTEL study-specific template. The four ROIs were combined into a single volumetric mask to generate a fifth ROI representing total BF volume. These five ROIs were used to extract GM volumetric data consisting of the sum of the total intensity of voxels within each ROI adjusted for the number of voxels. Each volume was corrected for head size using TIV as a proxy by applying a residual approach (i.e., residuals from linear regressions), as recommended by Wang et al. [45] see Ray et al., [9] for similar approach applied to BF volumes in PD].

### Statistical analyses

Demographic and clinical characteristics were compared between cognitive and VH status groups separately, using Mann-Whitney U (due to non-normality of some variables on the Kolmogorov-Smirnov test), and Chi-square tests. Rank-biserial correlations *r* and OR with 95%CI were reported as effect sizes. The relationships between the Total BF volume, VH and MCI were examined initially using Mann-Whitney U, followed by logistic regressions in which five covariates were added to account for potential confounds. These included age and sex, given their inclusion in previous research on the BF and MCI [9,12,46] and VH [7], as well as LEDD, DA (yes/no), and ACB (yes/no) given their potential contribution to MCI and VH symptoms in PD. Separate binary logistic regressions were conducted where the outcome variable was cognitive status (MCI/Intact cognition) or VH status (noVH/VH), followed by a multinomial regression where the outcome was a combination of these factors (i.e., MCI-VH, MCI-noVH, Intact cognition-noVH), excluding the rare Intact cognition-VH. From the latter analysis, three pairwise contrasts were examined. Statistical threshold for the Total BF variable was set at *p* <.05 corrected for five comparisons using the Benjamini-Hochberg FDR method. Following these main analyses, similar exploratory logistic regressions were conducted separately for specific BF ROIs (Ch1-2, Ch3, Ch4 and Ch4p). Statistical threshold for each ROI was set at *p* <.05 corrected for the four ROIs within each group comparison analysis using the FDR method. Logistic regression assumptions were examined using appropriate statistical tests (i.e., Variance Inflation Factors, Box-Tidwell procedure, Cook’s distance). Of note, our statistical approach deviates from our pre-registered plan due to sample characteristics, reviewers’ feedback, and statistical considerations. Detailed explanations are provided in Supplementary Materials.

### Data sharing

All data are part of the patients’ clinical charts, and patients have not provided consent to share their data publicly as this is a retrospective study.

## RESULTS

### Participant characteristics

Patient characteristics and related statistics are presented in Tables 1 and 2 according to cognitive status and VH status, and in Table S2 for the combination of symptoms. Overall, 61.1% of patients presented with PD-MCI and 25.3% with VH. These symptoms co-occurred in 20.6% of the sample, and few participants presented with VH without MCI (4.7%). There were no differences between cognitive or VH status groups for age, education, disease duration, LEDD, ACB, or MDS-UPDRS-III OFF scores. The VH group had a higher proportion of people taking DA than the noVH group. The MCI group had higher MDS-UPDRS-III ON scores and were less responsive to levodopa than the Intact cognition group, but did not differ with respect to DA.

**Table 1.** Characteristics of the PD-Intact Cognition and PD-MCI groups.

|  | Intact Cognition<br>n = 115 | MCI<br>n = 181 | Statistics |
| --- | --- | --- | --- |
| **Characteristics** |  |  |  |
| Female | 30.4% | 26.5% | $\chi^2 = 0.53, p = .47, OR = 0.83, CI [0.49-1.38]$ |
| Age (years) | 60.38 (7.77) | 61.75 (7.41) | $U = 9319.5, p = .13, r = -.09$ |
| Education (years) | 14.62 (3.19) | 14.12 (3.04) | $U = 9338.5, p = .13, r = -.09$ |
| Disease duration (years) | 9.24 (3.67) | 10.18 (4.25) | $U = 9257.5, p = .11, r = -.09$ |
| MDS-UPDRS-III ON | 14.54 (8.99) | 17.83 (10.05) | $U = 8090, p < .001, r = -.19$ |
| MDS-UPDRS-III OFF<sup>a</sup> | 37.45 (14.51) | 40.00 (14.46) | $U = 8644.5, p = .07, r = -.11$ |
| Levodopa response<sup>a</sup> | 62.40 (16.48) | 56.64 (16.63) | $U = 7754, p = .002, r = -.18$ |
| **Medications** |  |  |  |
| Total LEDD (mg) | 1345.75 (607.34) | 1430.70 (633.67) | $U = 11260, p = .24, r = -.07$ |
| DA (yes) | 53.0% | 48.1% | $\chi^2 = 0.70, p = .40, OR = 0.82, CI [0.51-1.31]$ |
| ACB $\geq 3$ | 12.2% | 18.2% | $\chi^2 = 1.93, p = .17, OR = 1.61, CI [0.82-3.16]$ |
| **Basal forebrain volumes (std)<sup>b</sup>** |  |  |  |
| Total BF | 0.25 (0.94) | -0.15 (1.00) | $U = 8070, p = .001, r = -.19$ |
| Ch1-2 | 0.23 (1.00) | -0.15 (0.97) | $U = 8364, p = .004, r = -.17$ |
| Ch3 | 0.18 (1.02) | -0.11 (0.97) | $U = 9019, p = .05, r = -.11$ |
| Ch4 | 0.22 (0.88) | -0.14 (1.04) | $U = 8229, p = .002, r = -.18$ |
| Ch4p | 0.09 (0.91) | -0.06 (1.05) | $U = 9453, p = .18, r = -.08$ |

*Note.* Data are presented as mean (SD), and statistics are uncorrected. MDS-UPDRS-III:
Movement Disorders Society—Unified Parkinson's Disease Rating Scale part 3; LEDD:
Levodopa Equivalent Daily Dose; DA: dopamine agonist; ACB: Anticholinergic Cognitive
Burden scale. Levodopa response = ((OFF–ON)/OFF\*100%). CI are 95%. <sup>a</sup>Seven missing
scores. <sup>b</sup> Standardized residuals corrected for total intracranial volume.

**Table 2.**
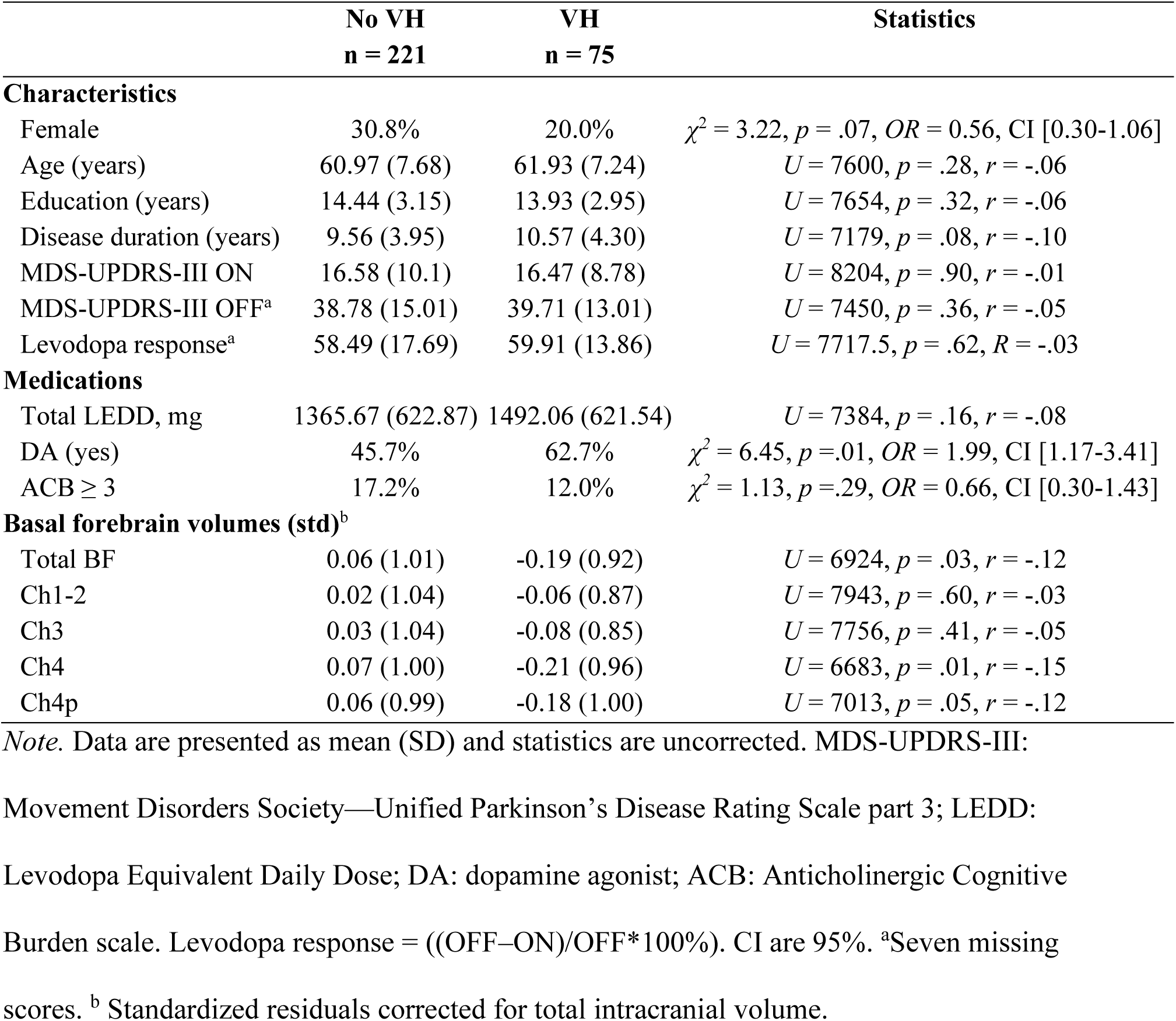
Characteristics of the PD-noVH and PD-VH groups.

### Total basal forebrain volume, MCI, and VH

Univariate group comparisons of BF volumes are presented in Tables 1, 2, and S2 (uncorrected). Given the potential contribution of confound variables (i.e., age, sex, LEDD, DA, and ACB), the relationships between Total BF volume and MCI, VH and their co-occurrence were assessed using logistic regressions. All models met statistical assumptions. OR with 95%CI are depicted in Figure 1a for Total BF volume and covariates. Results show Total BF volume to be negatively associated with the likelihood of MCI (*p* = .004; FDR-corr. *p* = .02), but not that of VH (*p* = .13). Null findings for VH status were replicated after excluding individuals without VH treated with antipsychotic medication (n=16; *p =* .12). Total BF volume was also negatively associated with the likelihood of co-occurring MCI and VH relative to neither symptom (*p* = .006; FDR-corr. *p* = .02). However, no significant relationship was observed in the likelihood of MCI in non-hallucinators (*p* = .08), or of VH in patients with MCI (*p* = .18). These results suggest that smaller BF volume is associated with MCI, but not VH when investigated in isolation. However, when considering both symptoms, smaller BF volume shows a modest relationship with the co-occurrence of MCI and VH relative to neither symptom. Despite the significant effect for MCI when VH is not considered, Total BF volume did not increase the likelihood of MCI in the absence of VH.

**Figure 1.**
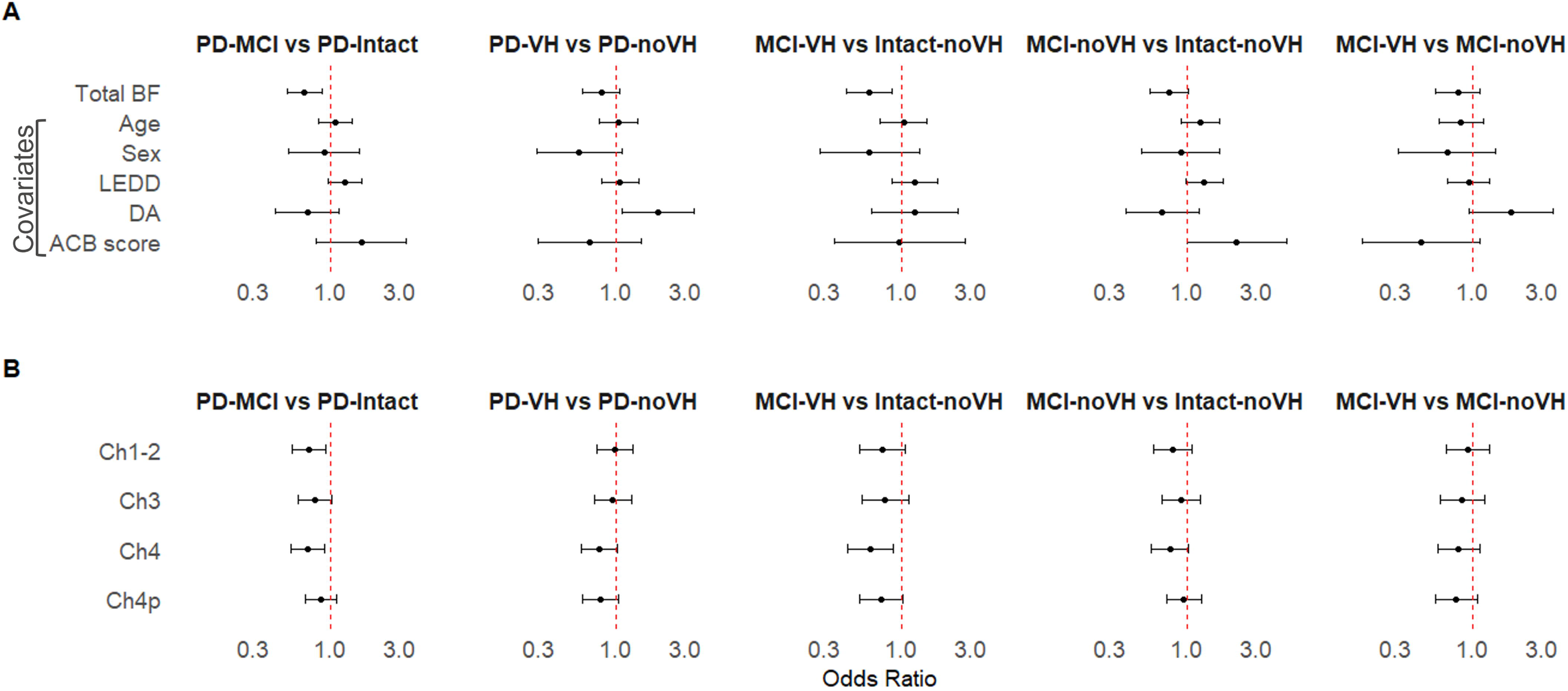
Comparisons of basal forebrain volumes across groups as a function of cognitive and VH status. *Note.* (a) OR and 95% CI of Total BF standardized residuals (corrected for TIV) after covarying age, sex, total levodopa equivalent daily dose (LEDD), dopamine agonist (DA, yes/no), and anticholinergic cognitive burden score (ACB≥3) in logistic regressions. (b) OR and 95% CI of regional BF volumes derived from independent logistic regressions after covarying age, sex, LEDD, DA, and ACB.

### Regional basal forebrain nuclei, MCI and VH

To examine the relationship between the BF, VH and MCI further, similar exploratory analyses were conducted with the four regional BF nuclei (Ch1-2, Ch3, Ch4, Ch4p). Univariate group comparisons of these volumes are presented in Tables 1, 2, and S2 (uncorrected), and separate logistic regressions accounting for potential confounds were conducted. These analyses met statistical assumptions except for Ch3 in analyses involving VH, and these results should be interpreted with caution. OR and 95%CI for each ROI across analyses are depicted in Figure 1b. In models investigating symptoms in isolation, both Ch1-2 and Ch4 volumes were negatively related to the likelihood of MCI (Ch1-2: *p* = .01; FDR-corr. *p* = .03; Ch4: *p* = .007; FDR-corr. *p* = .03). However, other nuclei did not show significant relationships with MCI (Ch3: *p* = .08; Ch4p: *p* = .26). In keeping with null findings obtained with the Total BF volume, regional BF volumes were not significantly associated with the likelihood of VH (*p ≥* .08). In models considering symptoms’ co-occurrence, only Ch4 volume showed a statistically significant relationship with MCI-VH relative to Intact cognition-noVH (*p* = 007; FDR-corr. *p* = .03). Volumes in other ROIs in this group contrast and all ROIs in other group contrasts did not reach statistical significance (*p ≥* .07, or FDR-corr. *p ≥* .14). Together, these results suggest that reduced Ch1-2 and Ch4 volumes relate to MCI when VH is not considered, and only reduced Ch4 volume relates significantly to the co-occurrence of MCI and VH relative to neither symptom.

## DISCUSSION

Our study is novel in that we examine the relationships between BF volumes, MCI and VH in a single cohort of individuals with long disease duration, and hence test the cholinergic phenotype hypothesis by bridging the literature linking the BF to each type of symptom separately. When examining symptom types separately and accounting for demographic variables and pharmacotherapy, we show that smaller Total BF volumes are modestly associated with higher odds of being diagnosed with MCI, but not VH. Smaller Ch4 and Ch1-2 are also modestly associated with higher odds of PD-MCI when VH symptoms are not considered. When both symptom types are considered, smaller Total BF and Ch4 volumes modestly relate to the co-occurrence of MCI and VH relative to neither symptom, and no other associations reached statistical threshold. Together, our results suggest that BF volumes, and particularly Ch4 volumes, are related to MCI, and this relationship is most robust when VH and MCI co-occur, which is consistent with the cholinergic phenotype hypothesis [4]. Ch1-2 volumes also appear to relate to MCI when VH status is not considered, which is interesting given its projection to temporal lobe regions, but this effect was not evident when considering both symptom types.

While we are first to examine both VH and MCI concurrently, our findings implicating the Total BF and Ch4 in MCI and in the co-occurrence of MCI and VH are consistent with the literature investigating these symptoms (in isolation). Although variability in findings across studies is noted, Total BF or Ch4 volume loss was previously shown to modestly relate to, or predict, each symptom in early PD [7–9] as well as performance on general cognition, memory, attention, executive functions, and visuospatial functions tests [9,12,46,47]. While the Total BF, including Ch4, does not exclusively comprise of cortically-projecting cholinergic neurons [43], recent evidence using more direct measurements of cortical acetylcholine (F-FEOBV PET) in PD demonstrates reductions in patients with cognitive decline [4], or in PD-VH [48]. Importantly, significant relationships were demonstrated between BF volumes and acetylcholine levels measured by PET across cortical and subcortical regions in PD [48,49], and BF volume was shown to mediate the relationship between acetylcholine levels and memory in the lateral neocortex and hippocampus [47]. Together, this evidence provides further support for the cholinergic hypothesis.

As for other BF nuclei, we found a modest relationship between Ch1-2 volume and the likelihood of PD-MCI, but found no significant effects for Ch3 or Ch4p. Only a few studies in PD report findings in these nuclei. One study showed Ch4p was not predictive of developing PD-MCI [12]. Studies investigating Ch1-2 or Ch1-2-3 volumes also yielded null findings when comparing PD-MCI or PD-VH to their cognitively intact or non-hallucinator counterparts [7,9] or PD patients to controls [46,49]. In contrast, one study found Ch1-2-3 volumes were smaller in *de novo* PD patients compared to controls and were correlated with visuospatial abilities and attention, but not memory [12]. The latter is surprising as the hippocampus is a major projection area of Ch1-2 and is critical to memory functioning. However, another study demonstrated a significant and specific relationship between Ch1-2 volume and visual memory in PD, which was mediated by hippocampal volumes [46]. The variability in findings between studies, including the present one, may be due to methodological differences in imaging analyses pipeline, diagnostic criteria, or patient characteristics including disease duration. Notably, volume loss in BF nuclei could follow a similar posterior to anterior pattern of progression as that seen in the Alzheimer’s disease spectrum [40], and thus Ch1-2 nuclei may be less affected at the early stages of PD.

Our study has several limitations, which may be addressed in future research. PD patients in our sample were being considered for DBS due to dyskinesia and motor fluctuations. Therefore, our findings may not generalize to the entire PD population, including patients who would be ineligible for DBS due to older age, presence of dementia, or severe balance problems. Despite this, the proportion of patients from our cohort presenting with concurrent VH and PD-MCI (21%) is similar to that reported in broader PD groups in a systematic review (28%) [1]. Furthermore, other studies investigating BF integrity in PD-MCI or as a predictor of future PD-VH leveraged data from a similar clinical group, albeit without accounting for medications in their statistical models [10,12].

Second, we relied on patient self-report during the clinical neuropsychology interview to ascertain the presence of VH and then coded these responses on a validated PD-specific scale. While we recognize that there are several standardized methods to assess VH in PD, they are not without limitations [50]. Gathering information from a semi-structured interview enables patients to describe their experience in their own words and allows clinicians to ask for elaboration. We also recognize that most patients with VH had minor hallucinations/illusions, and stronger relationships with BF volumes may be found in in more severe forms. However, findings showing relationships between Ch4 and future VH were also based on minor VH measured on the same MDS-UPDRS item [7].

Third, because MRI scans were acquired as part of the clinical care, variability in acquisition parameters may have introduced measurement error despite resampling to a common isometric voxel size. However, voxel size did not differ between groups, and thus, did not introduce a systematic bias (Table S4). Differences in results across regional BF may also reflect differences in the sizes of the original ROIs (Ch4>Ch1-2>Ch3=Ch4p), although measures were z-transformed. We deviated from our pre-registered analyses plan due to sample characteristics, statistical assumptions and reviewers’ suggestions (supplementary materials). We also cannot comment on the degree of atrophy given the absence of a healthy control group. Lastly, while VBM imaging methodology can provide insights based on group data, it is not amenable for use at the individual level. To our knowledge, no in-vivo technique to date provides the necessary precision to measure BF integrity for clinical use.

In conclusion, our findings support the notion that PD is a heterogeneous disease with individual differences not only in the degree of dopaminergic, but also of cholinergic dysfunction. Our study is in line with the cholinergic phenotype [4] as the co-occurrence of MCI and VH is associated with greater BF volume loss, particularly in Ch4. Future research is needed to examine the prevalence of the co-occurrence of other symptoms associated with this phenotype and relation to the integrity of the cholinergic system. This knowledge will contribute to the development of a unified method of assessing cholinergic compromise in PD, which would benefit the design of clinical trials targeting the cholinergic system and treatment planning in clinical care.

## Data Availability

All data being used for the study are part of the patients' clinical charts and therefore cannot be made public.

## ACKNOWLEDGMENTS

We thank Bianca Iddiols and Megan Vaziri for their help with data extraction and coding.

## AUTHORS’ ROLES

(1) Research project: A. Conception, B. Organization, C. Execution; (2) Statistical analysis: A. Design, B. Execution, C. Review and critique; (3) Manuscript preparation: A. Writing of the first draft, B. Review and critique.

S.A.: 1A, 1B, 1C, 2A, 2B, 3A, 3B

V.M.: 1C, 3B

M.S.: 1C, 2C, 3C

B.K.: 1C, 3B.

T.W.S.: 1A, 2C, 3B

A.C.R.: 1A, 2C, 3B

M.C.: 1A. 1B, 1C, 2A, 2B, 2C, 3A, 3B

## DISCLOSURES

### Funding Sources and Conflict of Interest

This work was supported by the Canadian Institutes of Health Research (CIHR; FRN 156447 & 162197) and UHN Collaborative Academic Practice. The authors declare that there are no conflicts of interest relevant to this work.

### Financial Disclosures for the previous 12 months

M.C. and M.S. received funding from the Michael J. Fox Foundation for Parkinson’s Research; S.A. receives funding from the University of Toronto and UHN; A.C.R. received funding from CIHR, SSHRC, the CAMH Foundation, and the University of Toronto, and received a honorarium from the University Institute of Mental Health in Montreal; T.W.S. received funding from the Alzheimer’s Society of Canada, CIHR, NSERC, Canada Foundation for Innovation, Western University, and the Ontario Ministry of Colleges and Universities; and BK and VM receive funding from CIHR.

### Ethical Compliance Statement

This study was approved by the Research Ethics Board at University Health Network. Informed consent was not necessary given the retrospective, clinical chart review design of the study. We confirm that we have read the Journal’s position on issues involved in ethical publication and affirm that this work is consistent with those guidelines.

## Notes

### Competing Interest Statement

The authors have declared no competing interest.

### Funding Statement

Canadian Institutes of Health Research (CIHR) and
UHN Collaborative Academic Practice.

### Author Declarations

The research ethics board of Toronto Western Hospital gave ethical approval for this work.

### Summary of Updates

The statistical model has been modified, however the main findings remain the same as the original submission.

