## Supplementary materials for "Basal forebrain volume and concurrent hallucinations and mild cognitive impairment in Parkinson’s disease"

### Table S1. Patients' characteristics across combined cognitive and VH statuses

| | Intact cognition | | MCI | | |
|---|---|---|---|---|---|
| | **No VH** | **VH** | **No VH** | **VH** | **Statistics** |
| | **n = 101** | **n = 14** | **n = 120** | **n = 61** | |
| **Characteristics** | | | | | |
| Female | 32.7% | 14.3% | 29.2% | 21.3% | $\chi^2$ (3) = 3.83, *p* = .28 |
| Age (years) | 59.89 (7.83) | 63.95 (6.49) | 61.89 (7.46) | 61.46 (7.36) | *H*(3) = 6.64, *p* = .08, *r* = .09 |
| Education (years) | 14.72 (3.17) | 13.86 (3.37) | 14.2 (3.14) | 13.95 (2.87) | *H*(3) = 2.87, *p* = .41, *r* = .06 |
| Disease Duration (years) | 9.17 (3.59) | 9.74 (4.34) | 9.88 (4.21) | 10.76 (4.30) | *H*(3) = 4.81, *p* = .19, *r* = .07 |
| MDS-UPDRS-III ON | 14.98 (9.39) | 11.36 (4.25) | 17.93 (10.52) | 17.97 (9.54) | *H*(3) = 12.07, *p* = .007, *r* = .12 |
| MDS-UPDRS-III OFF [a] | 37.71 (15.19) | 35.61 (8.53) | 39.66 (14.87) | 40.33 (14.01) | *H*(3) = 3.70, *p* = .30, *r* = .07 |
| Levodopa response [a] | 61.59 (17.16) | 68 (9.28) | 55.94 (17.79) | 55.42 (25.37) | *H*(3) = 11.95, *p* = .008, *r* = .12 |
| **Basal forebrain volumes (std)[b]** | | | | | |
| Total BF | 0.26 (0.99) | 0.20 (0.47) | -0.10 (1.02) | -0.28 (0.97) | *H*(3) = 13.11, *p* = .004, *r* = .12 |
| Ch1-2 | 0.20 (1.04) | 0.46 (0.59) | -0.13 (1.02) | -0.18 (0.88) | *H*(3) = 9.83, *p* = .02, *r* = .11 |
| Ch3 | 0.15 (1.03) | 0.39 (0.96) | -0.08 (1.05) | -0.18 (0.79) | *H*(3) = 5.87, *p* = .12, *r* = .08 |
| Ch4 | 0.25 (0.92) | 0.04 (0.50) | -0.08 (1.05) | -0.27 (1.05) | *H*(3) = 12.60, *p* = .006, *r* = .12 |
| Ch4p | 0.11 (0.94) | -0.05 (0.65) | 0.02 (1.03) | -0.21 (1.07) | *H*(3) = 4.95, *p* = .18, *r* = .07 |
| **Medications** | | | | | |
| Total LEDD (mg) | 1315.51 (606.86) | 1563.91 (585.95) | 1407.90 (635.48) | 1475.57 (632.91) | *H*(3) = 3.56, *p* = .31, *r* = .06 |
| DA (yes) | 48.5% | 85.7% | 43.3% | 57.4% | $\chi^2$ (3) = 10.69, *p* = .01 |
| Specific DA drugs | | | | | |
| Bromocriptine | - | 7.1% | - | - | |
| Pramipexole | 35.6% | 35.7% | 29.2% | 36.1% | |
| Ropinorole | 5.9% | 35.7% | 9.2% | 19.7% | |
| Rotigotine | 6.9% | 7.1% | 6.7% | 1.6% | |
| ACB ≥ 3 | 11.9% | 14.3% | 21.7% | 11.5% | $\chi^2$ (3) = 2.83, *p* = .42 |
| Drugs with ACB Score = 3 | | | | | |
| Amitriptyline | 1.0% | - | - | 1.6% | |
| Atropine | - | - | 0.8% | - | |
| Clozapine[c] | - | - | 0.8% | - | |
| Dimenhydrinate | - | - | 1.7% | - | |
| Diphenhydramine | 1.0% | - | - | - | |
| Fesoterodine | - | | 0.8% | 1.6% | |
| Methocarbamol | 1.0% | 7.1% | - | - | |
| Olanzapine[c] | - | - | 1.7% | - | |
| Oxybutynin | - | - | - | 1.6% | |
| Paroxetine | 1.0% | 7.1% | - | - | |
| Quetiapine[c] | 5.0% | 7.1% | 5.8% | 13.3% | |
| Solifenacin | 1.0% | - | - | - | |
| Tolterodine | 1.0% | - | 2.5% | 1.6% | |

| | | | | |
|---|---|---|---|---|
| Trihexyphenidyl | 3.0% | - | 5.8% | 4.9% |
| Drugs with ACB score = 2 | | | | |
| Amantadine | 30.7% | 42.9% | 35.8% | 41.0% |
| Cyclobenzaprine | - | - | 0.8% | - |
| Drugs with ACB score = 1 | | | | |
| Alprazolam | 1.0% | - | - | - |
| Bupropion | 1.0% | - | 1.7% | - |
| Fentanyl | - | - | - | 1.6% |
| Venlafaxine | 3.0% | - | 2.5% | - |
| Cognitive enhancers | | | | |
| Donepezil | 1.0% | - | 0.8% | - |
| Modafinil | - | - | - | 1.6% |

*Note*: Data are presented as mean (SD), and statistics are uncorrected. MDS-UPDRS-III: Movement Disorders Society—unified Parkinson's Disease Rating Scale part 3; LEDD: Levodopa Equivalent Daily Dose; DA: dopamine agonist; ACB: Anticholinergic Cognitive Burden scale. Levodopa response = ((OFF–ON)/OFF*100%); MCI: Mild Cognitive Impairment; VH: Visual Hallucinations. [a] Seven missing scores. [b] Standardized residuals corrected for total intracranial volume. [c] antipsychotic medications.

Chi-squares and Kruskal Wallis H tests were used to compare groups. The only significant differences (uncorrected) between the four groups were seen in the motor severity score in the ON state, levodopa response and DA.

**Table S2. Visual hallucination severity and types**

| | **Intact-noVH** **n =14** | **MCI-VH** **N=61** |
|---|---|---|
| VH severity score (1:2:3) | 79% : 21% : 0% | 79%: 18% : 3% |
| VH score 1: subtypes | n = 11 | n = 48 |
| Illusions/mispercep. only | 27% | 29% |
| Presence/passage only | 55% | 42% |
| Both types | 18% | 29% |

**Table S3. MCI subtypes and cognitive domains affected**

| | **MCI-noVH** **n = 120** | **MCI-VH** **n = 61** | **Statistics** |
|---|---|---|---|
| MCI multiple domains | 87.5% | 91.8% | $\chi^2$ = 0.76, *p* = .38, *OR* = 1.60, 95%CI [.55-4.63] |
| Cognitive domains: | | | |
| Executive | 78.3% | 90.2% | $\chi^2$ = 3.89, *p* = .05, *OR* = 2.54, 95%CI [.98-6.54] |
| Attention/speed | 70.0% | 80.3% | $\chi^2$ = 1.90, *p* = .17, *OR* = 1.68, 95%CI [.80-3.54] |
| Memory | 45.0% | 62.3% | $\chi^2$ = 4.84, *p* = .03, *OR* = 2.02, 95%CI [1.08-3.79] |
| Visuoperception/spatial | 44.2% | 49.2% | $\chi^2$ = 0.41, *p* = .52, *OR* = 1.22, 95%CI [.66-2.27] |
| Language | 35.8% | 29.5% | $\chi^2$ = 0.72, *p* = .40, *OR* = 0.75, 95%CI [.39-1.46] |

In terms of cognitive domains, memory was more likely to be impaired in the MCI-VH than in the MCI-no VH group based on chi-squared tests, although this would not survive corrections of multiple comparisons.

**Table S4. MRI acquisition parameters**

| Voxel size at acquisition (x*y*z) | n | Number of Slices (range) |
|---|---|---|
| 0.39 x 0.39 x 1.00 | 90 | 124 to 182 |
| 0.41 x .041 x 1.00 | 11 | 140 to 170 |
| 0.43 x 0.43 x 1.00 | 29 | 144 to 196 |
| 0.45 x 0.45 x 1.00 | 3 | 158 to 166 |
| 0.47 x 0.47 x 1.00 | 7 | 166 to 186 |
| 0.49 x 0.49 x 1.00 | 1 | 186 |
| 0.51 x 0.51 x 1.00 | 1 | 162 |
| 0.86 x 0.86 x 1.00 | 138 | 142 to 178 |
| 0.90 x 0.90 x 1.00 | 6 | 156 to 166 |
| 0.94 x 0.94 x 1.00 | 9 | 146 to 166 |
| 1.00 x 1.00 x 1.00 | 1 | 156 |

All T1-weighted spoiled gradient recalled echo (SPGR) structural MRI scans were acquired on the same 3T Signa MR system (GE Medical Systems, Milwaukee, WI). Voxel size acquisition varied as these scans were collected as part of clinical care. All scans were resliced to a 1x1x1mm voxel dimensions prior to other pre-processing steps. Of note, voxel size did not differ between groups based on MCI status ($U$ = 10385, $p$ = .97; $r$ = .00) or VH status ($U$ = 8144, $p$ = .81; $r$ = -.01), suggesting that there was no systematic bias. Furthermore, only small correlations are noted with standardized BF volumes (Total BF: $r_s$ = .00, $p$ = .95; Ch1-2: $r_s$ = .19, $p$ <.001; Ch3: $r_s$ = .13, $p$ = .02; Ch4: $r_s$ = -.09, $p$ = .13; Ch4p: $r_s$ = -.18, $p$ = .002).

**Transparency and openness**

The present study has been preregistered (https://doi.org/10.17605/OSF.IO/QAW6G). There were some deviations from the preregistered statistical plan due to data characteristics, reviewers' comments, and error in logic related to covariates in the selection of the statistical plan. First, we planned to test main effects of MCI, VH and their interaction jointly. However, this was not appropriate given the uneven case distributions (Table S1), and specifically, given the rare occurrence of VH in the context of intact cognition. Therefore, main effects were tested separately on the full sample in two analyses, and the effect of the co-occurrence of MCI and VH (i.e., interaction) was tested after creating a combined variable with three levels (Intact cognition-noVH; MCI-noVH, and MCI-VH) and excluding participants presenting with Intact cognition-VH (n=14).

Second, we used logistic regressions with diagnoses as outcome variables instead of the proposed ANCOVAs in which BF volumes were the dependant variables. This was motivated as we realized our initial plan included an important error in logic related to covariates. Specifically, the covariates included are thought to relate mainly to diagnostic categories (e.g., MCI or VH) rather than BF volumes. For instance, medications are thought to worsen MCI and VH symptoms. As such, they are theoretically appropriate to use when the outcome variables are diagnoses (MCI, VH or combination) rather than when the outcome variable is BF volume (ANCOVAs).

In addition, the total BF volume, extracted from a mask combining Ch1-4p, was added prior to analyzing subregional BF volumes separately, due to collinearity and spatial overlap of the regional ROIs. The DA variable was binarized (yes/no) instead of reflecting dosage because many patients did not take DA, a covariate reflecting anticholinergic cognitive burden was added given that these medications can worsen cognition, and analyses involving VH were repeated after excluding individuals without VH who were prescribed antipsychotic medications.
